# Performance verification of detecting COVID-19 specific antibody by using four chemiluminescence immunoassay systems

**DOI:** 10.1101/2020.04.27.20074849

**Authors:** Yafang Wan, Zhijie Li, Kun Wang, Tian Li, Pu Liao

**Affiliations:** Department of Clinical Laboratory, Chongqing General Hospital, Chongqing, China

**Keywords:** COVID-19, Antibody, Chemiluminescence immunoassay, Performance verification

## Abstract

**Background:** The purpose of current study is to evaluate the analytical performance of seven kits for detecting IgM/IgG antibody of corona virus (2019-nCoV) by using four chemiluminescence immunoassay systems.

**Methods:** 50 patients diagnosed with 2019-nCoV infection and 130 controls without corona virus infection from the General Hospital of Chongqing were enrolled in current retrospective study. Four chemiluminescence immunoassay systems including seven IgM/IgG antibody detection Kits for 2019-nCoV (A_IgM, A_IgG, B_IgM, B_IgG, C_IgM, C_IgG, D_Ab) were employed to detecting antibody concentration. Chi-square test, receiver operating characteristic (ROC) curve and Youden’s index were demonstrated to verify the cutoff value of each detection system.

**Results:** The repeatability verification results of the A, B, C, and D system are all qualified. D-Ab performances best (92% sensitivity and 99.23% specificity), and B_IgM worse than other systems. Except for the system of A_IgM and C_IgG, the optimal diagnostic thresholds and cutoff value of other kits from recommendations are inconsistent with each other. B_IgM got the worst AUC and C_IgG had the best diagnostic accuracy. More importantly, B_IgG system have the highest false positive rate for testing patients with AIDS, tumor and pregnant. A_IgM system test showed highest false positive rates among elder over 90 years old.

**Conclusions:** Systems for CoVID-2019 IgM/IgG antibody test performance difference. Serum diagnosis kit of D-Ab is the most reliable detecting system for 2019-nCoV antibody, which can be used as an alternative method for nucleic acid testing.

## Introduction

The corona virus pneumonia (Corona Virus Disease 2019, COVID-19) is an acute respiratory infection caused by severe acute respiratory syndrome coronavirus type 2 (SARS-CoV-2) [1]. The epidemic of the disease has not ended since the winter of 2019 and it is still raging around the world. SARS-CoV-2 is highly contagious through air, droplets and contacts. Generally, the incubation period of SARS-CoV-2 is 3-7 days, but the longest incubation period can reach 14 days [2]. It has caused more than 2,420,000 people infections and nearly 167,000 deaths worldwide until the end of April 21th. Therefore, the early diagnosis of SARS-CoV-2 infection is very crucial. Previous studies show that SARS-CoV-2 antigen stimulates the immune system to produce an immune response, and specific IgM and IgG antibodies will appear in the serum of patients after infecting [3]. The SARS-CoV-2 specific IgM and IgG antibody tests have been involved in the diagnosis criteria for suspected cases whose COVID-19 viral nucleic acid test appears false negative in recently published guideline of Novel Corona Virus Pneumonia Diagnosis and Treatment (Trial Version 7) which advocated by the National Health Committee [4].

Current popularly detection methods for SARS-CoV-2 antibodies include colloidal gold and chemiluminescence immunoassay [5]. Chemiluminescence immunoassay is a laboratory technology that combines a luminescence system with an immune response. It not only has the specificity of the immune response, but also has the high sensitivity of the luminescence reaction, and is widely used in immunoassays [6]. Our laboratory currently has four automatic chemiluminescence immunoassay systems A, B, C and D, of which the three detection systems A, B and C detect SARS-CoV-2 specific IgM and IgG antibodies respectively, and the D system detects total antibody of IgM/IgG. Current investigation intends to evaluate the repeatability, clinical sensitivity and specificity of 7 antibody detection kits for 4 detecting systems, as well as the false positive rate in special populations. The Youden’s index verifies the best diagnostic threshold (Cutoff value) of each detection system to understand the analytical performance of each system detecting and ensure the detecting results.

## Material and Methods

### Sample Collection

50 serum samples from patients with SARS-CoV-2 diagnosed in January 2020 and 130 serum samples from patients with other conditions including 20 late pregnancy women, 20 patients with solid tumors, 20 patients with AIDS, 21 patients over 90 years old and 49 normal controls were enrolled from the Immunology Department of the Laboratory Department of Chongqing General Hospital (The Third Hospital) from late February to March 2020. All collected serum specimens are inactivated in a water bath at 56 ℃ for 1 hour, and then stored in a refrigerator at -80℃.

### Reagents and instruments

Automatic immunochemiluminescence analyzer A called detection system A (Bioscience Diagnostic Technology Co., Ltd.). Reagents include the Coronavirus (2019-nCoV) IgM antibody detection kit (Referred to A_IgM, batch number: G202002415), and S/CO (Sample CutOff value) ≥ 1.0 denoted to be positive. 2019-nCoV IgG antibody detection kit (Referred to A_IgG, batch number: G202002414), and S/CO ≥ 1.0 denoted to be positive. Fully automatic immunochemiluminescence analyzer B called detection system B (Shenzhen New Industries Biomedical Engineering Co., Ltd.). Reagents include the 2019-nCoV IgM antibody detection kit (Referred to B_IgM, batch number: 271200201), and S/CO≥1.0 AU/ml denoted to be positive. 2019-nCoV IgG antibody detection kit (Referred to B_IgG, batch number: 2722000101), and S/CO≥1.0 AU/ml denoted to be positive. Automatic immunochemiluminescence analyzer C called detection system C (Shenzhen YHLO Biotech Co., Ltd.). Reagents include the severe acute respiratory syndrome coronavirus 2 (SARS-CoV-2) IgM antibody detection kit (Referred to C_IgM, batch number: 20200206), and S/CO≥10 AU/ml denoted to be positive. SARS-CoV-2 IgG antibody detection kit (Referred to C_IgG, batch number: 20200202), and S/CO ≥ 10 AU/ml denoted to be positive. Fully automatic immunochemiluminescence analyzer D called detection system D (Xiamen Innodx Biotech Co., Ltd.). Reagents include the 2019-nCoV antibody detection kit (Referred to D_Ab, batch number: 20200309), and S/CO≥1.0 denoted to be positive.

### Precision verification

Under the condition of calibration and quality control of the detection systems, all of them are qualified and the following experiments are carried out.

The cutoff value (cutoff value) is 1.0, 1.0, 1.10 AU/ml, 1.10 AU/ml, 10 AU/ml, 10 AU/ml and 1.0 in detection system of A_IgM, A_IgG, B_IgM, B_IgG, C_IgM, C_IgG, D_Ab respectively. In 50 specimens of patients infected SARS-CoV-2, one case of weak positive specimen with S/CO value within less than 3 times of cutoff value (Level1, L1) and one case with an S/CO value greater than 3 times of cutoff value (Level 2, L2) were selected. Within-run precision was conducted firstly. All detecting system analyzes their corresponding L1 and L2 specimens respectively, conducting 20 consecutive tests. All test were completed within one day, observe 20 S/CO value, judge the result, and calculate its standard deviation and coefficient of variation. The result is judged to be 100% in line, and the coefficient of variation is less than 10% is qualified. Between-run precision was conducted secondly. Detecting system analyze the corresponding L1 and L2 specimens once a day, and continuously detecting for 20 days, observation 20 times S/CO value, judge the result, and calculate its standard deviation and coefficient of variation. The result is judged to be 100% in line, and the coefficient of variation is less than 15% is qualified.

### Statistical analysis

All statistical analyses were conducted using R software (http://www.R-project.org/). Evaluation of sensitivity with 95% CI, specificity with 95% CI and false positives in specific populations were conducted separately. Use ROC curve (R packages pROC) and Youden’s index to calculate the optimal diagnostic threshold (Cutoff value) of the detection system.

## Results

In order to test precision of each kit, we performed within-run and between-run detecting. As can be seen from **Table 1**, the repeatability verification results of the A, B, C, and D system are all qualified. Among them, system D performance best and systems B performance worst in the weak positive specimens. More important, system of B_IgM and B_IgG are nearly twice as precise as C_IgM and D-Ab.

**Table 1.**
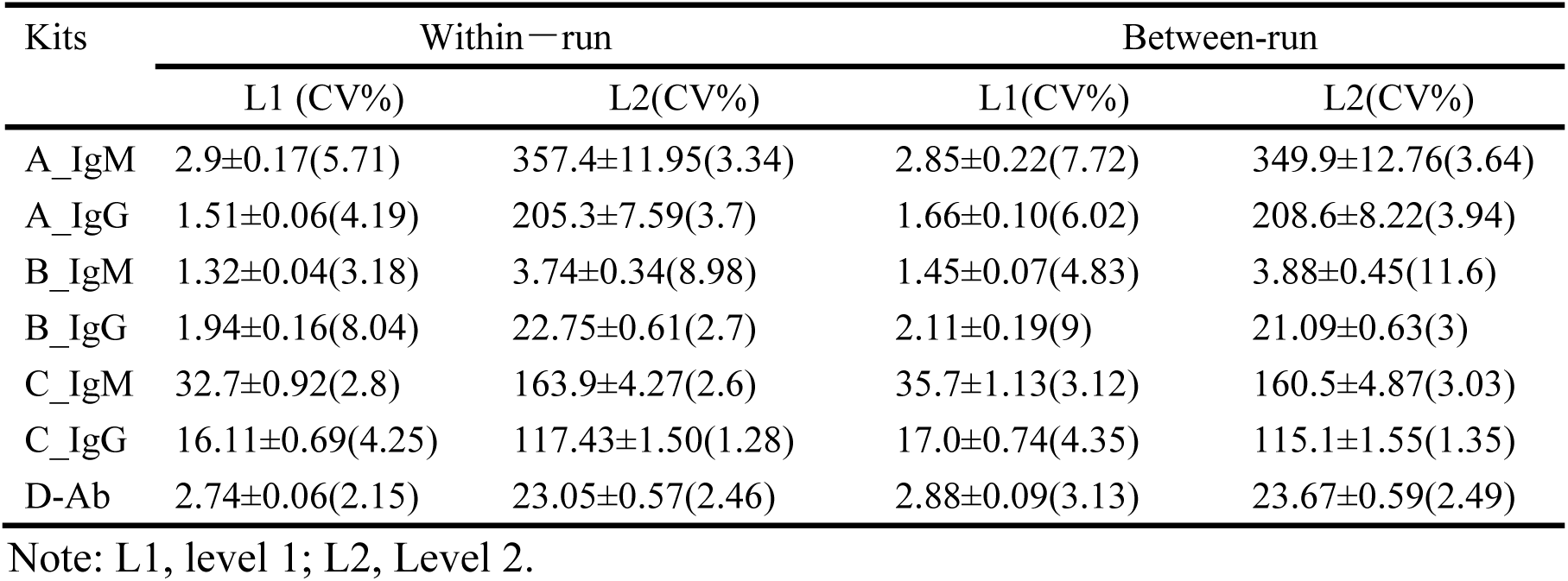
Diagnosis precision within different Kits

Total 50 patients were considered as COVID-19 because viral nucleic acid test appears positive, and other 130 controls got negative result of viral nucleic acid. Overall 180 subjects were tested for COVID-19 specific serological assay. The results showed vary sensitivity and specificity among different kits. D-Ab performances best (92% sensitivity and 99.23% specificity), and B_IgM worse than others (**Table 2**).

**Table 2.** Diagnosis sensitivity and specificity within different Kits

| Kits | Positive in Case | Positive in Control | Sensitivity [95%CI] | Specificity [95%CI] |
| --- | --- | --- | --- | --- |
| A_IgM | 41(9) | 8(122) | 82.00% [69.20%,90.23%] | 93.85% [88.33%,96.85%] |
| A_IgG | 43(7) | 4(126) | 86.00% [73.81%,93.05%] | 96.92% [92.36%,98.80%] |
| B_IgM | 13(37) | 8(122) | 26.00% [15.87%,39.55%] | 93.85% [88.33%,96.85%] |
| B_IgG | 43(7) | 28(102) | 86.00% [73.81%,93.05%] | 78.46% [70.63%,84.66%] |
| C_IgM | 31(19) | 3(127) | 62.00% [48.15%,74.14%] | 97.69% [93.44%,99.21%] |
| C_IgG | 44(6) | 3(127) | 88.00% [76.20%,94.38%] | 97.69% [93.44%,99.21%] |
| D-Ab | 46(4) | 1(129) | 92.00% [81.16%,96.85%] | 99.23% [95.77%,99.86%] |

ROC curve was depicted by using original S/CO value (Figure1). According to ROC curve, we can get optimal operating point of different kits (**Table 3**). It can be conclude that, except for the optimal operating thresholds of A_IgM and C_IgG, the optimal diagnostic thresholds of other kits and the Cutoff value from recommendations are inconsistent with each other. The results showed that the AUC of D_Ab reached 0.95 and Youden’s index is 0.93 (**Table 3**). The optimal cutoff value was 0.54, with sensitivity and specificity values of 99% and 94%, respectively. According to the optimal operating threshold, there were only 3 patients who had a negative result and two controls had a positive result. Totally, B_IgM got the worst AUC and C_IgG had the best diagnostic accuracy.

**Table 3.** Cutoff value and ROC related parameters within different Kits

| Kit | Cutoff Value | Optimal Operating Point | Specificity | Sensitivity | AUC | Youden's index |
| --- | --- | --- | --- | --- | --- | --- |
| A-IgM | 1 | 0.9 | 0.96 | 0.82 | 0.94 | 0.78 |
| A-IgG | 1 | 0.49 | 0.95 | 0.9 | 0.95 | 0.85 |
| B-IgM | 0.9 | 0.56 | 0.82 | 0.44 | 0.61 | 0.26 |
| B-IgG | 0.9 | 3.17 | 0.99 | 0.78 | 0.89 | 0.77 |
| C-IgM | 10 | 1.85 | 0.84 | 0.96 | 0.95 | 0.8 |
| C-IgG | 10 | 9.61 | 0.98 | 0.9 | 0.96 | 0.88 |
| D-Ab | 1 | 0.54 | 0.99 | 0.94 | 0.95 | 0.93 |

**Figure1.**
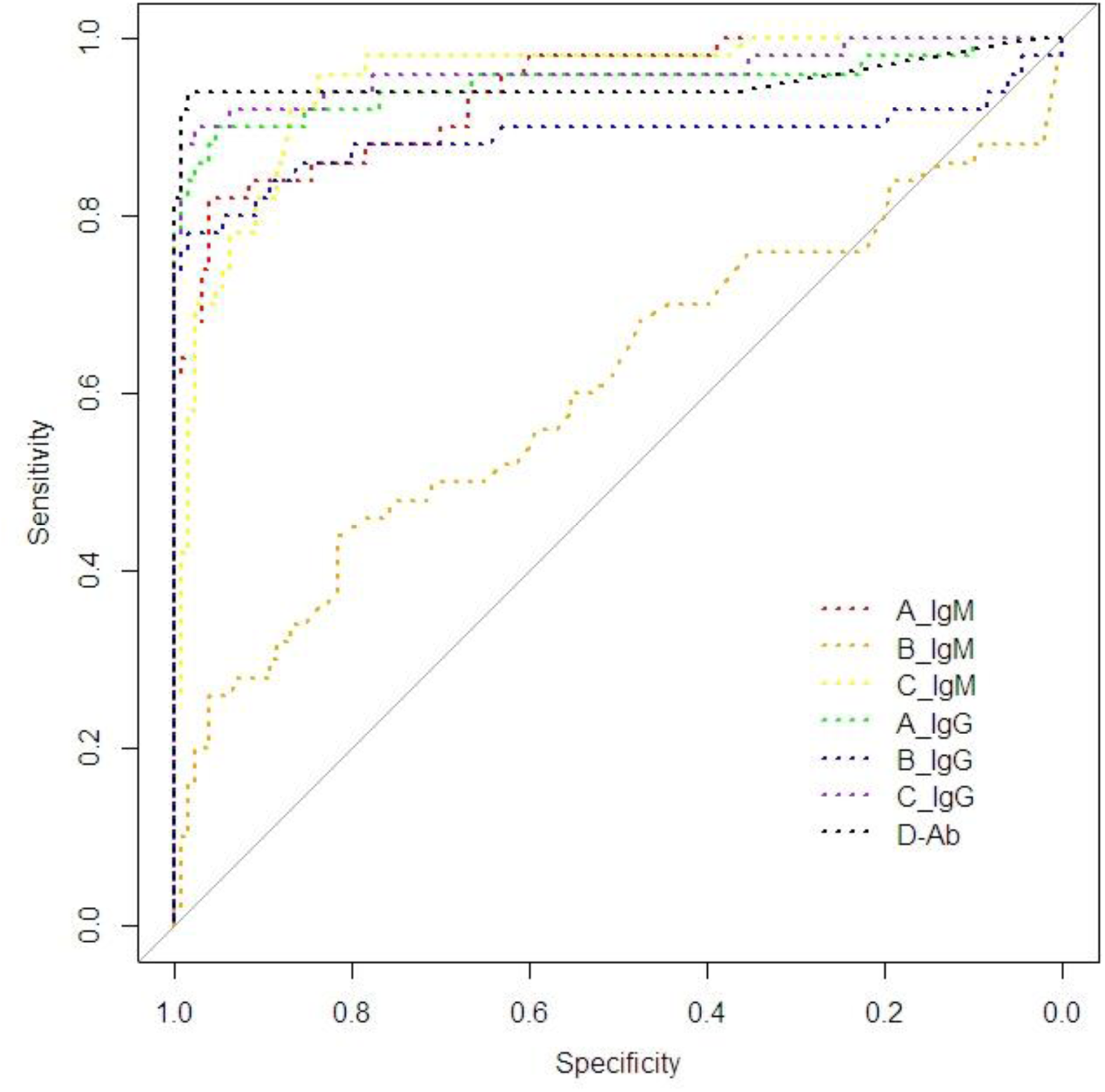
ROC curve within different Kits

Considering endogenous and exogenous factors exist in the process of antibody assay, subgroups of controls including patients with AIDS (Acquired Immune Deficiency Syndrome), tumor, pregnant and elders more than 90 years old were involved in current analysis. Each system has false positive results in the selected subgroup of controls (**Table 4**). It is worth noting that B_IgG system have the highest false positive rate for testing patients with AIDS, tumor and pregnant. A_IgM system test showed the highest false positive rates among elder over 90 years old.

**Table 4.** The false positive rate in specific patients

| Sub_group | A-IgM | A-IgG | B-IgM | B-IgG | C-IgM | C-IgG | D-Ab |
| --- | --- | --- | --- | --- | --- | --- | --- |
| HIVs | 5 | 0 | 5 | 55 | 5 | 5 | 5 |
| Tumors | 10 | 15 | 10 | 40 | 5 | 0 | 0 |
| Pregnants | 5 | 0 | 10 | 15 | 0 | 0 | 0 |
| Elder(≥90) | 9.5 | 0 | 4.8 | 0 | 4.8 | 4.8 | 0 |

## Discussions

SARS-CoV-2 belongs to the β genus and is the seventh well-known corona virus infected human beings. Its nucleocapsid protein (NP) stimulates human immune system to cause chemical reactions. Specific IgM antibodies emergence at the 7th day of infecting, and appear at peaks after 28 days. Specific IgG antibody emerged around the 10th day of infecting, and reached peaks after 49 days, which can maintain at a long time in the blood. The median time for total plasma antibodies appear at the 12th day after infecting [7, 8]. In current investigation, the average time of serum collection in all subjects at 13 days after diagnosis, therefore it is considered that specific IgM and IgG antibodies should already exist in the specimen.

With the published guideline of Novel Coronavirus Pneumonia Diagnosis and Treatment Program (Trial Version 7) [4], the suspected cases comply with condition of positive for serum specific 2019-nCoV IgM/IgG antibodies, or the specific 2019-nCoV IgG antibody performance positive, or 2019-nCoV IgG antibody is 4 times in recovery period than that of the acute period can confirm diagnosis of COVID-19 [9, 10]. The diagnosis standard of COVID-19 changed the situation. There is a huge market demand for SARS-CoV-2 antibody detection reagents worldwide. Manufacturers domestically produce antibody detection reagents which are used in the clinical laboratory. Previous investigation has shown that the clinical specificity and sensitivity of some 2019-nCoV IgM antibody are 96.2% and 70.24%, respectively. And the clinical specificity and sensitivity of 2019-nCoV IgG antibody are 92.4% and 96.1% [6]. Therefore, the false negative and false positive results will appear in the detection, which will cause confusion to clinical judgment. So the laboratory needs to pay close attention to the performance indicators of the reagents used.

Seven detection kits from four chemiluminescence systems were used in current study. All of the kits have been permitted to use in the emergency approval of the China National Drug Administration or the EU CE sales, and have been applied to clinical detecting. According to the requirements of People's Republic of China Health Industry Standard WS/T 505-2017 “Qualitative Measurement Performance Evaluation Guidelines” [10], the performance indicators of qualitative kits should focus on repeatability, clinical accuracy (including clinical sensitivity and specificity) and verification of Cutoff value. The results showed that the repeatability of all detecting systems is in line with the manufacturer's statement, but the variance among them is relatively big. Specifically, the coefficients of variation regarding B-IgM and B-IgG are larger than others.

According to guideline of WS/T 494-2017, the sensitivity and specificity of qualitative items for different occasions are also regulated. In the using of preliminary screening tests, the sensitivity should be greater than 95%. In the occasion of diagnose, both of the sensitivity and specificity should be greater than 95%. In a confirmed diagnostic test, the specificity should be greater than 98% [9]. According to the result of current study, the clinical sensitivity and specificity from all detecting systems does not meet the requirements of screening, diagnosis and confirm diagnosis experiments. Therefore, all detection systems cannot be used independently for the diagnosis and of SARS-CoV-2 infections, and need to be used together with nucleic acids test and clinical symptoms considering.

Regarding the confounding factors influence detection results, we divided controls for subgroups which includes patients with AIDS, tumor, pregnant and older people over 90 years old [5,11–12]. The results of current investigation show that B_IgM has the lowest sensitivity, indicating the possibility of higher false negatives which can occur at the window period of viral infection or at the patients with lower immunity.

B_IgG has the lowest specificity which indicated higher false positives and prone to occur for special patients, such as AIDS, solid tumor, pregnant and the elderly, etc.[13] The reason of false positives may be due to some interfering substances (such as rheumatoid factor, homologous to the kit antibodies, etc.) present in the specimen. Simultaneously, according to the area under the ROC curve of each detecting system, it is found that the diagnostic accuracy of B_IgM and B_IgG also demonstrated worst, and the diagnostic accuracy of the other systems is better. In addition, according to ROC curve and Youden’s index, the best diagnostic thresholds exist in A-IgM and C-IgG, and others are inconsistent with the manufacturer's declaration. The optimal threshold of A_IgG, B_IgM, C_IgM, D_Ab are less than the Cutoff value indicating more false positive results. The optimal threshold of C_IgM is greater than the Cutoff value, indicating more false negative cases.

Therefore, the laboratory should conduct the necessary performance evaluation of the selected novel coronavirus antibody, carefully interpret the results of the novel coronavirus antibody, make the necessary further testing requirements, and reduce the missed diagnosis and misdiagnosis.

## Data Availability

Relevant study data will be made available on reasonable request from the corresponding author at

## Acknowledgements

The project was supported partly by grants from National Natural Science Foundation of China (81572089), and partly by grants from Yuzhong District Scientific Research Project (20180129).

## Author Contributions

Y.W. and P.L. conceived the project. Y.W., Z.L. and K.W. were responsible for the tissue sample collection. Y.W., Z.L. and T.L. performed the laboratory test. Y.W. conducted the statistical analysis. Y.W. and P.L. wrote the manuscript, and responsible for all data present in current research.

## Ethics approval and consent to participate

The current investigation was conducted with the approval of the medical ethics committee of the General Hospital of Chongqing.

## Data available

Relevant study data will be made available on reasonable request from the corresponding author at

